# Comparison of Leadless and Transvenous Pacemakers in Post-TAVR Patients: A Large-Scale Real-World Analysis

**DOI:** 10.64898/2025.12.03.25341596

**Authors:** Dylan Pereira, Thibault Lenormand, Carl Semaan, Alexandre Bodin, Laurent Fauchier, Arnaud Bisson

## Abstract

**Background:** Permanent pacemaker implantation (PPI) is required in up to 30% of patients following transcatheter aortic valve replacement (TAVR), mainly due to conduction disturbances. While transvenous pacemakers (TVPM) have traditionally been used, leadless pacemakers (LLPM) could improve outcomes and reduce complications, especially in high-risk populations.

**Methods:** We conducted a retrospective, observational, propensity-matched cohort study using the TriNetX Global Collaborative Network. Patients who underwent TAVR followed by single chamber PPI within 30 days were identified and divided into two groups: LLPM and TVPM. Those with pre-existing pacemakers or concomitant cardiac surgery were excluded. Propensity score matching (1:1) was performed on demographic, clinical, laboratory, echocardiographic, and medication variables. Clinical outcomes such as all-cause mortality, heart failure, atrial fibrillation, procedural and device-related complications during follow-up were assessed. Kaplan–Meier analyses and Cox proportional hazards models were used.

**Results:** Among 1,425 identified patients (469 LLPM, 956 TVPM), 367 matched pairs were analyzed. At 5-year follow-up, there was no significant difference in all-cause mortality (27.2% LLPM vs. 29.4% TVPM; HR 1.07, 95% CI 0.81–1.40; p=0.637). LLPM was associated with a significantly lower incidence of heart failure (16.1% vs. 25.6%; HR 0.61, 95% CI 0.44–0.84; p=0.002). Atrial fibrillation during follow-up was lower in the LLPM group (18.0% vs. 25.6%; HR 0.69, 95% CI 0.50–0.95; p=0.018). Procedure and device-related complication rates were low and similar between groups.

**Conclusions:** In this large real-world cohort, LLPM was associated with a reduced risk of heart failure and atrial fibrillation compared to TVPM, without differences in overall mortality or safety. These data support considering leadless pacing in selected post-TAVR patients, although prospective randomized studies are required to confirm these findings.

**Clinical Perspective:** *What Is Known:*

- Permanent pacemaker implantation is frequent after transcatheter aortic valve replacement (TAVR) and may negatively impact outcomes through device related complications, pacing-induced dyssynchrony and tricuspid interference.
- Leadless pacemakers eliminate transvenous leads and pockets, reducing the risk of lead-related and infectious complications.

*What the Study Adds:*

- In this large real-world propensity-matched analysis, leadless pacemakers were associated with fewer heart failure and atrial fibrillation events compared with transvenous devices, without differences in mortality or procedural safety.
- These findings support the use of leadless pacing as a safe and potentially morbidity-sparing option in selected post-TAVR patients.

## Introduction

Transcatheter aortic valve replacement (TAVR) has become the standard of care for patients with severe symptomatic aortic stenosis who are at high or intermediate surgical risk.^1–4^ While this procedure has significantly improved patient outcomes, it remains associated with a high incidence of conduction disturbances, leading to permanent pacemaker implantation (PPI) in approximately 9–30% of cases.^5–8^ These conduction abnormalities are primarily caused by mechanical compression or direct injury to the conduction system, particularly at the level of the aortic annulus, due to its anatomical proximity to the bundle of His and left bundle branch.^5,8,9^ These patients often exhibit high long-term mortality, frequently from non-cardiovascular causes.^10^ Moreover, multiple studies have shown that pacemaker implantation following TAVR may adversely impact prognosis^11,12^, particularly due to pacing-induced dyssynchrony and valve-device interactions.

Traditionally, transvenous pacemakers (TVPMs) have been the first-line treatment for post-TAVR conduction disorders.^5,8^ However, their use is associated with various complications, including lead displacement, venous obstruction, and an increased risk of infection.^13–15^ These concerns have led to the development of leadless pacemakers (LLPMs), which eliminate the need for transvenous leads and a subcutaneous pocket, potentially reducing device-related complications.^16–22^

Recent studies have suggested that LLPMs may offer safety advantages over TVPMs, particularly in reducing long-term lead-related issues.^6,23,24^ Large-scale registry data, including findings from the Micra Coverage with Evidence Development (CED) study, indicate that LLPMs are associated with a lower incidence of chronic complications and device-related reinterventions.^5,20,21,24^ Additionally, emerging evidence suggests that LLPMs may be particularly beneficial in high-risk populations, such as patients with chronic kidney disease, malignancy, or other comorbidities.^21,25^ Despite these findings, real-world data comparing the clinical outcomes of LLPMs and TVPMs in post-TAVR patients remain limited.^23^

This study aims to evaluate the real-world outcomes of LLPM vs. TVPM in patients requiring PPI after TAVR, leveraging data from the TriNetX Global Collaborative Network. The primary objective is to assess short- and mid-term outcomes, including all-cause mortality, atrial fibrillation, heart failure, endocarditis, and major complications.

## Patients and Methods

### Patient selection

This was a retrospective, observational, and exploratory cohort study using data from the TriNetX Global Collaborative Network, a federated health research platform providing access to electronic health records (EHRs) from approximately 300 million de-identified patients across 147 large healthcare organizations (HCOs) in 17 countries worldwide.

This analysis compared outcomes in patients who required PPI within 30 days post-TAVR, with one group receiving a leadless pacemaker (LLPM) and the other a single chamber transvenous pacemaker (TVPM) between January 2015 and December 2024. These procedures were identified based on the presence of respective International Classification of Disease-10th Revision Procedure Coding System codes in any position (Appendix A). Patients with a pre-existing permanent pacemaker or defibrillator were excluded.

The analysis process included two main steps: 1) Defining the cohorts through query criteria; 2) Setting up and running the analysis. Setting up the analysis required definitions for the index event, outcomes criteria, and the time frame. This query was run on the network Global Collaborative Network with 147 HCO(s) queried and 147 HCO(s) responded. A total of 45 provider(s) responded with patients. The follow-up of patients was up to 5 years after inclusion.

### Collected data and propensity score matching

The baseline parameters collected are presented in **Table 1**. These included demographics, race and ethnicity, body mass index (BMI), blood pressure, and cardiovascular comorbidities such as hypertension, diabetes mellitus, smoking status, overweight and obesity, dyslipidemia, heart failure, coronary artery disease, dilated cardiomyopathy, valvular heart disease, amyloidosis, arrhythmias, and conduction disorders. Non-cardiovascular comorbidities included, among others, lung disease, kidney disease, a history of nicotine dependence, alcohol abuse, thyroid disease, anemia, sleep apnea syndrome, and a history of malignancy.

**Table 1.** Baseline characteristics of patients before and after propensity score matching. Values are expressed as mean ± SD for continuous variables and n (%) for categorical variables. * Unreliable due to small cell size <10

|  | Before propensity-score matching |  |  |  | After propensity-score matching |  |  |  |
| --- | --- | --- | --- | --- | --- | --- | --- | --- |
|  | LLPM<br>(n = 469) | TVPM<br>(n = 956) | P-Value | Std<br>diff. | LLPM<br>(n = 367) | TVPM<br>(n = 367) | P-Value | Std<br>diff. |
| **Demographics** |  |  |  |  |  |  |  |  |
| Age | 79.1 ± 8.3 | 79.3 ± 7.6 | 0.703 | 0.021 | 79.9 ± 7.5 | 79.9 ± 7.5 | 0.941 | 0.005 |
| Men | 262 (55.9) | 554 (57.9) | 0.454 | 0.042 | 202 (55.0) | 204 (55.6) | 0.882 | 0.011 |
| Body mass index (kg/m2) | 28.8 ± 6.8 | 28.9 ± 6.4 | 0.808 | 0.015 | 28.9 ± 6.9 | 29.0 ± 6.8 | 0.759 | 0.025 |
| White | 371 (79.1) | 782 (81.8) | 0.224 | 0.068 | 303 (82.6) | 304 (82.8) | 0.922 | 0.007 |
| Black or African American | 37 (7.9) | 38 (4.0) | 0.002 | 0.166 | 19 (5.2) | 21 (5.7) | 0.745 | 0.024 |
| **Comorbidities** |  |  |  |  |  |  |  |  |
| Hypertension | 365 (77.8) | 722 (75.5) | 0.337 | 0.054 | 286 (77.9) | 289 (78.7) | 0.788 | 0.020 |
| Systolic BP (mm Hg) | 120.5 ±<br>26.6 | 121.5 ±<br>27.6 | 0.530 | 0.039 | 121.6 ±<br>25.4 | 122.4 ±<br>25.9 | 0.726 | 0.028 |
| Diastolic BP (mm Hg) | 61.4 ± 16.1 | 60.6 ± 15.4 | 0.384 | 0.054 | 62.3 ± 14.7 | 61.1 ± 14.5 | 0.309 | 0.082 |
| Diabetes mellitus | 212 (45.2) | 378 (39.5) | 0.041 | 0.115 | 152 (41.4) | 153 (41.7) | 0.940 | 0.006 |
| Overweight or obesity | 143 (30.5) | 280 (29.3) | 0.641 | 0.026 | 104 (28.3) | 106 (28.9) | 0.870 | 0.012 |
| Malnutrition | 51 (10.9) | 59 (6.2) | 0.002 | 0.169 | 31 (8.4) | 30 (8.2) | 0.894 | 0.010 |
| Dyslipidaemia | 374 (79.7) | 710 (74.3) | 0.023 | 0.130 | 287 (78.2) | 298 (81.2) | 0.313 | 0.075 |
| Heart failure | 354 (75.5) | 580 (60.7) | < 0.001 | 0.322 | 262 (71.4) | 263 (71.7) | 0.935 | 0.006 |
| Coronary artery disease | 379 (80.8) | 753 (78.8) | 0.370 | 0.051 | 294 (80.1) | 302 (82.3) | 0.450 | 0.056 |
| Acute myocardial infarction | 73 (15.6) | 110 (11.5) | 0.031 | 0.119 | 47 (12.8) | 55 (15.0) | 0.393 | 0.063 |
| Dilated cardiomyopathy | 13 (2.8) | 21 (2.2) | 0.054 | 0.037 | ≤ 10 | ≤ 10 | 1.000 | < 0.001* |
| Amyloidosis | ≤ 10 | 12 (1.3) | 0.207* | 0.068 | ≤ 10 | ≤ 10 | 1.000 | < 0.001* |
| Nonrheumatic mitral valve stenosis | 22 (4.7) | 66 (6.9) | 0.103 | 0.095 | 19 (5.2) | 25 (6.8) | 0.351 | 0.059 |
| Nonrheumatic mitral valve insufficiency | 109 (23.2) | 303 (31.7) | 0.001 | 0.190 | 89 (24.3) | 87 (23.7) | 0.863 | 0.013 |
| Nonrheumatic aortic valve stenosis | 462 (98.5) | 925 (96.8) | 0.054 | 0.115 | 361 (98.4) | 365 (99.5) | 0.155 | 0.105 |
| Nonrheumatic aortic valve insufficiency | 137 (29.2) | 270 (28.2) | 0.704 | 0.021 | 98 (26.7) | 111 (30.2) | 0.288 | 0.079 |
| Nonrheumatic aortic valve stenosis + insufficiency | 140 (29.9) | 357 (37.3) | 0.005 | 0.159 | 113 (30.8) | 123 (33.5) | 0.429 | 0.058 |
| Rheumatic mitral valve diseases | 43 (9.2) | 53 (5.5) | 0.010 | 0.139 | 30 (8.2) | 33 (9.0) | 0.693 | 0.029 |
| Ischemic stroke | 56 (11.9) | 81 (8.5) | 0.037 | 0.115 | 43 (11.7) | 40 (10.9) | 0.727 | 0.026 |
| Intracranial hemorrhage | ≤ 10 | ≤ 10 | 0.101* | 0.087 | ≤ 10 | ≤ 10 | 1.000 | <<br>0.001* |
| Atrial fibrillation or flutter | 270 (57.6) | 509 (53.2) | 0.123 | 0.087 | 213 (58.0) | 207 (56.4) | 0.654 | 0.033 |
| Persistent atrial fibrillation | 102 (21.7) | 152 (15.9) | 0.007 | 0.150 | 81 (22.1) | 77 (21.0) | 0.719 | 0.027 |
| Chronic atrial fibrillation | 145 (30.9) | 278 (29.1) | 0.476 | 0.040 | 122 (33.2) | 121 (33.0) | 0.937 | 0.006 |
| Sick sinus syndrome | 44 (9.4) | 49 (5.1) | 0.002 | 0.165 | 32 (8.7) | 34 (9.3) | 0.796 | 0.019 |
| Atrioventricular and LBB block | 340 (72.5) | 442 (46.2) | < 0.001 | 0.555 | 245 (66.8) | 250 (68.1) | 0.694 | 0.029 |
| LBB block | 124 (26.4) | 175 (18.3) | < 0.001 | 0.196 | 89 (24.3) | 89 (24.3) | 1.000 | < 0.001 |
| Other and unspecified RBB block | 127 (27.1) | 169 (17.7) | < 0.001 | 0.227 | 87 (23.7) | 90 (24.5) | 0.796 | 0.019 |
| Acute kidney failure and chronic kidney disease | 239 (51.0) | 406 (42.5) | 0.002 | 0.171 | 160 (43.6) | 174 (47.4) | 0.299 | 0.077 |
| End stage renal disease | 83 (17.7) | 52 (2.3) | < 0.001 | 0.390 | 35 (9.5) | 36 (9.8) | 0.901 | 0.009 |
| Dependence on renal dialysis | 42 (9.0) | 47 (4.9) | 0.003 | 0.159 | 28 (7.6) | 29 (7.9) | 0.890 | 0.010 |
| Lung disease | 312 (66.5) | 556 (58.2) | 0.002 | 0.173 | 233 (63.5) | 241 (65.7) | 0.537 | 0.046 |
| Sleep apnea syndrome | 122 (26.0) | 188 (19.7) | 0.006 | 0.152 | 82 (22.3) | 85 (23.2) | 0.792 | 0.019 |
| Peripheral vascular disease | 83 (17.7) | 133 (13.9) | 0.061 | 0.104 | 56 (15.2) | 60 (16.2) | 0.686 | 0.030 |
| Neoplasms | 167 (35.6) | 286 (29.9) | 0.030 | 0.121 | 122 (33.2) | 119 (32.4) | 0.814 | 0.017 |
| Anemia | 180 (38.4) | 243 (25.4) | < 0.001 | 0.281 | 121 (33.0) | 123 (33.5) | 0.875 | 0.012 |
| Acute and subacute endocarditis | ≤ 10 | 11 (1.2) | 0.148* | 0.077 | ≤ 10 | ≤ 10 | 1.000 | <<br>0.001* |
| Other sepsis | 51 (10.9) | 64 (6.7) | 0.006 | 0.148 | 26 (7.1) | 29 (7.9) | 0.674 | 0.031 |
| Disorders of thyroid gland | 144 (30.7) | 246 (25.7) | 0.048 | 0.111 | 108 (29.4) | 106 (28.9) | 0.871 | 0.012 |
| Alzheimer's disease | ≤ 10 | 12 (1.3) | 0.207* | 0.068 | ≤ 10 | ≤ 10 | 1.000 | <<br>0.001* |
| Anxiety, stress-related and other mental disorders | 102 (21.7) | 149 (15.6) | 0.004 | 0.159 | 69 (18.8) | 80 (21.8) | 0.313 | 0.075 |
| Alcohol abuse | ≤ 10 | 16 (1.7) | 0.543* | 0.034 | ≤ 10 | ≤ 10 | 1.000 | <<br>0.001* |
| Nicotine dependence | 131 (27.9) | 295 (30.9) | 0.257 | 0.064 | 112 (30.5) | 136 (37.1) | 0.061 | 0.139 |
| Mood disorders | 93 (19.8) | 128 (13.4) | 0.002 | 0.174 | 63 (17.2) | 66 (18.0) | 0.771 | 0.021 |

| Laboratory and echocardiographic parameters |  |  |  |  |  |  |  |  |
| --- | --- | --- | --- | --- | --- | --- | --- | --- |
| Estimated GFR (MDRD, ml/min) | 54.3 ± 31.3 | 60.5 ± 26.2 | <0.001 | 0.215 | 59.4 ± 29.9 | 58.8 ± 25.5 | 0.802 | 0.019 |
| CRP (mg/L) | 37.9 ± 63.1 | 34.0 ± 61.5 | 0.622 | 0.062 | 36.2 ± 64.3 | 34.0 ± 52.4 | 0.821 | 0.038 |
| Hemoglobin (g/dl) | 11.1 ± 2.3 | 11.6 ± 2.3 | < 0.001 | 0.218 | 11.3 ± 2.2 | 11.3 ± 2.2 | 0.819 | 0.018 |
| LVEF (%) | 62.0 ± 8.8 | 56.7 ± 13.6 | 0.002 | 0.470 | 61.9 ± 9.0 | 59.1 ± 10.3 | 0.105 | 0.287 |
| Baseline treatments |  |  |  |  |  |  |  |  |
| Beta Blockers | 327 (69.7) | 602 (63) | 0.012 | 0.143 | 246 (67.0) | 250 (68.1) | 0.752 | 0.023 |
| ACE Inhibitors | 143 (30.5) | 247 (27.4) | 0.064 | 0.104 | 109 (29.7) | 101 (27.5) | 0.514 | 0.048 |
| Angiotensin II Inhibitors | 158 (33.7) | 262 (27.4) | 0.015 | 0.137 | 111 (30.2) | 119 (32.4) | 0.524 | 0.047 |
| Diuretics | 318 (67.8) | 607 (63.5) | 0.109 | 0.091 | 249 (67.8) | 246 (67.0) | 0.813 | 0.017 |
| SGLT2 inhibitors | 34 (7.2) | 41 (4.3) | 0.019 | 0.127 | 24 (6.5) | 24 (6.5) | 1.000 | < 0.001 |
| Drugs used in diabetes | 240 (51.2) | 381 (39.9) | < 0.001 | 0.229 | 173 (47.1) | 168 (45.8) | 0.711 | 0.027 |
| Antiplatelet therapy | 412 (87.8) | 684 (71.5) | < 0.001 | 0.414 | 312 (85.0) | 318 (86.6) | 0.525 | 0.047 |
| Rivaroxaban | 41 (8.7) | 84 (8.8) | 0.978 | 0.002 | 39 (10.6) | 35 (9.5) | 0.624 | 0.036 |
| Dabigatran | $\leq 10$ | 18 (1.9) | 0.750* | 0.018 | $\leq 10$ | $\leq 10$ | 1.000 | <<br>0.001* |
| Edoxaban | $\leq 10$ | $\leq 10$ | 0.101* | 0.087 | $\leq 10$ | $\leq 10$ | 1.000 | <<br>0.001* |
| Warfarin | 85 (18.1) | 184 (19.2) | 0.611 | 0.029 | 66 (18.0) | 60 (16.3) | 0.557 | 0.043 |
| Apixaban | 133 (28.4) | 207 (21.7) | 0.005 | 0.155 | 102 (27.8) | 99 (27.0) | 0.804 | 0.018 |

Laboratory and echocardiographic parameters included estimated glomerular filtration rate (eGFR) using the creatinine-based MDRD formula, C-reactive protein (CRP), hemoglobin, and left ventricular ejection fraction (LVEF).

Medication data were collected, including the presence (but not the dose) of beta-blockers, angiotensin-converting enzyme inhibitors, angiotensin receptor blockers, diuretics, SGLT2 inhibitors, diabetes medications, anticoagulants, and antiplatelet agents.

Propensity score matching was performed using all collected parameters for the entire study population.

### Study endpoint

The study endpoints included in-hospital outcomes as well as midterm (up to 5 years) outcomes, including all-cause death, heart failure hospitalization, atrial fibrillation, tricuspid regurgitation, infective endocarditis, and device-related complications. The in-hospital outcomes included various in-hospital complications, including overall complications, embolism and thrombosis, cardiac effusion/perforation/tamponade, puncture site complications, device-related complications (including mechanical breakdown, device dislodgement, other mechanical complication, infection and inflammatory reaction, hemorrhage, pain, stenosis caused by a prosthetic device, and pocket complication), other complications (including postprocedural hematoma or hemorrhage, intraoperative cardiac arrest, pericarditis, vascular complications, hemothorax, and pneumothorax), and in-hospital death.

The ICD-10-CM codes used to ascertain endpoints are summarized in Appendix C. Identification of heart failure hospitalization and atrial fibrillation during the follow-up period was performed using ICD-10-CM codes in the primary diagnosis upon rehospitalization subsequent to the procedure, thereby ensuring that it constituted the main reason for hospital admission rather than historical diagnoses.

The date of pacemaker implantation marked the beginning of the observation, with censoring occurring at the time of the event of interest, death, loss of follow-up within facilities involved in the TriNetX Global Collaborative Network, or up to 5 years after the procedure as applicable.

### Statistical analyses and outcome measures

Continuous variables are expressed as means with standard deviation (SD) or medians with interquartile ranges (IQR), and categorical variables are presented as absolute values and percentages. Comparisons between groups were performed using the appropriate statistical tests depending on the data distribution: t-test or Wilcoxon rank-sum test for continuous variables, and Chi-square or Fisher’s exact test for categorical variables.

Time-to-event outcomes were analyzed using Kaplan–Meier survival curves. The log-rank test was applied to compare survival distributions between groups when the proportional hazards assumption was met visually and statistically. In cases where the proportional hazards assumption was violated (e.g., crossing curves), the p-value derived from the test of proportionality was reported.

Hazard ratios (HRs) and 95% confidence intervals (CIs) were calculated for time-to-event outcomes using Cox proportional hazards models. For binary outcomes without time-to-event data or when survival analysis was not applicable, odds ratios (ORs) were reported.

A propensity score matching (PSM) approach was used to control for baseline differences between groups. Matching was performed on a large number of demographic, clinical, laboratory, and medication variables using a 1:1 greedy nearest neighbor algorithm without replacement and a caliper of 0.01. Standardized differences were calculated to assess post-matching balance.

To comply with TrinetX data privacy policies, any data point involving fewer than 10 patients per cell was automatically masked and displayed as “≤ 10”. This suppression rule may impact the precision of derived proportions and statistical comparisons in rare outcomes.

All analyses were conducted within the TriNetX platform using its integrated analytics tools. Statistical significance was set at p < 0.05.

## Results

### Baseline characteristics before and after propensity matching in the whole population

A total of 1,425 patients who underwent transcatheter aortic valve replacement (TAVR) and received a permanent pacemaker within 30 days of the procedure were identified in the TriNetX database. Among them, 469 patients received a leadless pacemaker (LLPM), and 956 received a single chamber transvenous pacemaker (TVPM). The study flowchart illustrating patient selection and cohort matching is shown in **Supplemental Figure S2.**

Baseline clinical, biological, echocardiographic, and therapeutic characteristics differed significantly between the two groups prior to matching, as summarized in **Table 1**. Patients receiving LLPMs were slightly younger, more frequently male, and had more comorbidities such as heart failure, atrial fibrillation, chronic kidney disease, and diabetes than patients implanted with TVPMs. They also received more cardiovascular medications including beta-blockers and oral anticoagulants.

A 1:1 propensity score matching was performed using all collected baseline variables, resulting in two matched cohorts of 367 patients each. After matching, the baseline characteristics were well balanced between the LLPM and TVPM groups, with most standardized mean differences below 0.1, indicating appropriate covariate balancing (**Figure S1**). The matched population was then used for all subsequent comparative analyses.

### Clinical outcomes

In the matched population, the mean follow-up was 626±558 days (median 448 [772]) for the LLPM group and 730±578 days (median 563 [924]) in the TVPM group; the IQR is reported as width (Q3–Q1) as provided by TriNetX. All-cause mortality occurred in 100 patients (27.2%) in the LLPM group and 108 patients (29.4%) in the TVPM group. The incidence of death over the 5-year follow-up did not significantly differ between the two groups (hazard ratio [HR] 1.07, 95% confidence interval [CI] 0.81–1.40; p = 0.637).

Heart failure was significantly less frequent in patients with LLPM compared to those with TVPM (59 [16.1%] vs. 94 [25.6%], HR 0.61, 95% CI 0.44–0.84; p = 0.002).

Endocarditis occurred in 47 patients (12.8%) in the LLPM group versus 42 patients (11.4%) in the TVPM group. The difference was not statistically significant (HR 1.23, 95% CI 0.81–1.86; p = 0.319).

Atrial fibrillation was observed in 66 patients (18.0%) in the LLPM group and 94 patients (25.6%) in the TVPM group, with a significantly lower incidence in the LLPM group (HR 0.69, 95% CI 0.50–0.95; p = 0.018).

Tricuspid regurgitation was present in 32 patients (8.7%) in the LLPM group and 47 patients (12.8%) in the TVPM group. Although this difference did not reach statistical significance, a trend toward a lower prevalence was observed in the LLPM group (HR 0.69, 95% CI 0.44–1.07; p = 0.098).

These results are summarized in **Figure 1** and **Table 2**.

**Figure 1.**
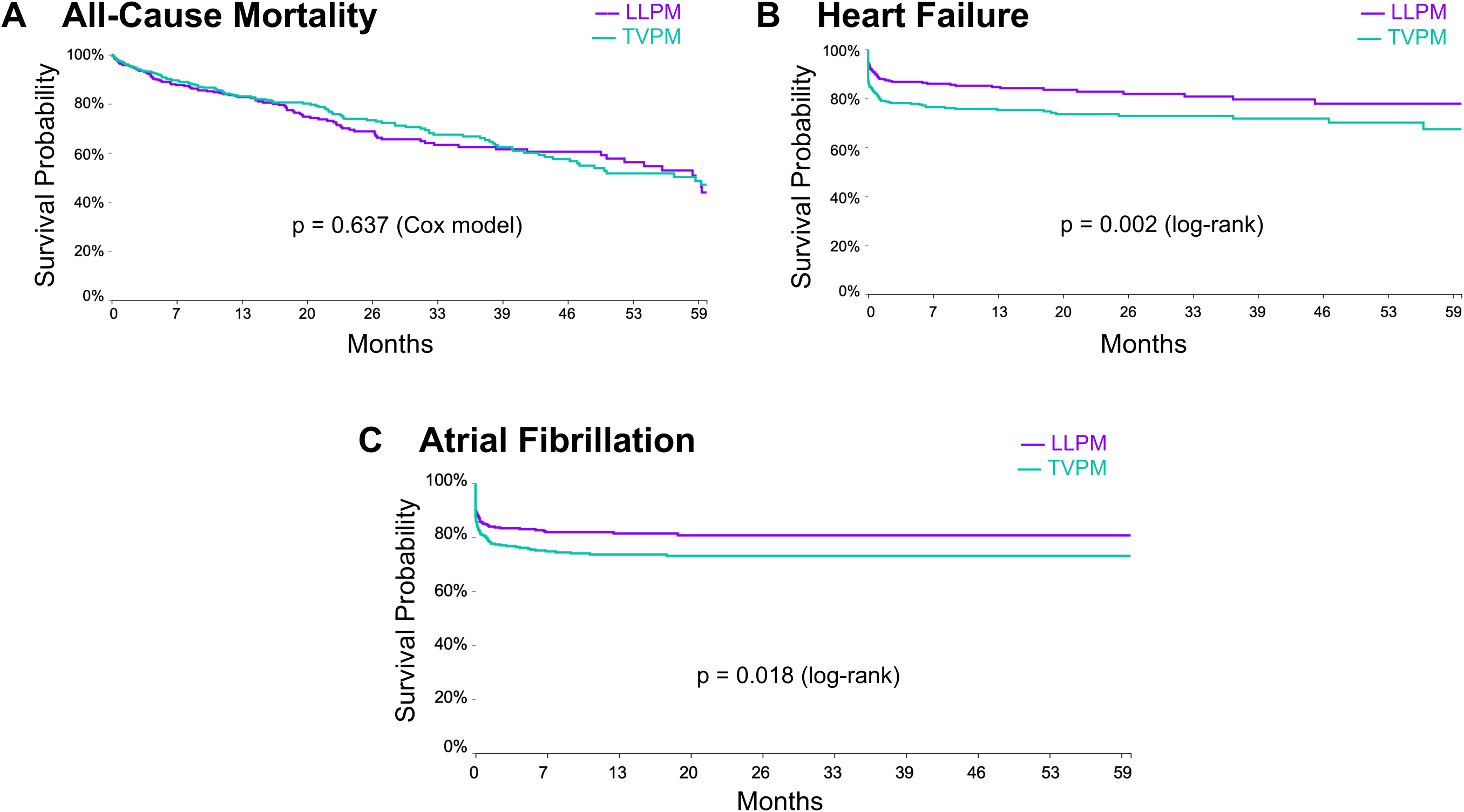
Kaplan–Meier curves for all-cause mortality (A), heart failure (B), and atrial fibrillation (C) after pacemaker implantation post-TAVR in matched LLPM and TVPM cohorts (up to 5 years)

**Table 2.** Clinical outcomes during follow-up after pacemaker implantation post-TAVR in matched LLPM and TVPM cohorts. Values are expressed as n (%). Hazard ratios (HR) are provided with 95% confidence intervals (CI). Follow-up duration was up to 5 years. LLPM: leadless pacemaker; TVPM: transvenous pacemaker.

|  | **LLPM** | **TVPM** |  |  |
| --- | --- | --- | --- | --- |
|  | **(n = 367)** | **(n = 367)** |  |  |
|  | **Number of events** | **Number of events** | **Hazard ratio**<br>**(95% CI)** | **p value** |
| Death | 100 (27.2) | 108 (29.4) | 1.069 (0.814, 1.403) | 0.637 |
| Heart failure | 59 (16.1) | 94 (25.6) | 0.606 (0.438, 0.839) | 0.002 |
| Atrial fibrillation | 66 (18.0) | 94 (25.6) | 0.690 (0.504, 0.946) | 0.018 |
| Tricuspid regurgitation | 32 (8.7) | 47 (12.8) | 0.685 (0.437, 1.074) | 0.098 |
| Endocarditis | 47 (12.8) | 42 (11.4) | 1.225 (0.808, 1.859) | 0.319 |

### Procedure-related complications

The composite outcome of overall complications (excluding heart failure and endocarditis) occurred in 33 patients (9.0%) in the LLPM group and 46 patients (12.5%) in the TVPM group (odds ratio [OR] 0.70, 95% CI 0.43–1.14; p = 0.150), without reaching statistical significance. Procedure-related complications occurred at low but comparable rates in both groups. Cardiac effusion or perforation was observed in 16 patients (4.4%) in the LLPM group and 18 patients (4.9%) in the TVPM group, without a statistically significant difference (OR 0.87, 95% CI 0.44–1.74; p = 0.692).

Puncture site complications were reported in ≤10 patients (≤2.7%) in the LLPM group and in 11 patients (3.0%) in the TVPM group (OR 0.90, 95% CI 0.38–2.14; p = 0.804). Similarly, post-procedural hematoma was observed in 0 patients in the LLPM group and in ≤10 patients (≤2.7%) in the TVPM group. Due to low event counts, no odds ratio could be calculated for this outcome, but the difference in risk was statistically significant (p = 0.001).

Pericarditis occurred in ≤10 patients (≤2.7%) in both groups (OR 1.00, 95% CI 0.41–2.43; p = 0.634), and intraoperative cardiac arrest was also rare (≤10 patients in both groups, HR 0.99, 95% CI 0.41–2.42; p = 0.990). Vascular complications were identified in ≤10 patients in each group. These results confirm that the safety profile of LLPM implantation was comparable to that of TVPM in terms of acute procedural risks. Full details are presented in **Table 3**.

**Table 3.** Complications observed during follow-up after pacemaker implantation post-TAVR in matched LLPM and TVPM cohorts. Values are expressed as n (%). Odds ratios (OR) are provided with 95% confidence intervals (CI). Follow-up duration was up to 5 years. HF: heart failure; IE: infective endocarditis. * p-value derived from risk difference comparison using z-test. ** No odds ratio could be calculated due to zero events in one group. p-value based on risk difference (z-test). “–“ : Not available or not calculated ^a^ Cells with 10 or fewer patients are reported as “≤ 10” in accordance with TrinetX data de-identification policy, which suppresses small cell counts to protect patient confidentiality.

|  | **LLPM** | **TVPM** |  |  |
| --- | --- | --- | --- | --- |
|  | **(n = 367)** | **(n = 367)** |  |  |
|  | **Number of events** | **Number of events** | **Odds ratio**<br>**(95% CI)** | **p value** |
| **Cardiac complications** |  |  |  |  |
| Overall complications (except HF & IE) | 33 (9.0) | 46 (12.5) | 0.702 (0.434, 1.137) | 0.150 |
| Cardiac effusion or perforation | 16 (4.4) | 18 (4.9) | 0.870 (0.436, 1.736) | 0.692* |
| Pericarditis | $\leq 10^a$ | $\leq 10^a$ | 1.000 (0.411, 2.432) | 0.634 |
| Intraoperative cardiac arrest | $\leq 10^a$ | $\leq 10^a$ | 0.994 (0.409, 2.419) | 0.990 |
| Device related complications | $\leq 10^a$ | 20 (5.4) | 0.491 (0.227, 1.065) | 0.067 |
| Other complications | $\leq 10^a$ | $\leq 10^a$ | 1.012 (0.416, 2.464) | 0.979 |
| **Vascular access related complications** |  |  |  |  |
| Puncture site complication | $\leq 10^a$ | 11 (3.0) | 0.896 (0.375, 2.137) | 0.804* |
| Vascular complication | $\leq 10^a$ | $\leq 10^a$ | 1.000 (0.411, 2.433) | 1.000 |
| Post procedural hematoma | 0 | $\leq 10^a$ | — | 0.001** |
| Post procedural haemorrhage | $\leq 10^a$ | $\leq 10^a$ | 0.997 (0.410, 2.426) | 0.995 |
| **Embolic and thrombotic complications** |  |  |  |  |
| Venous embolism and thrombosis | 32 (8.7) | 23 (6.3) | 1.429 (0.819, 2.492) | 0.099 |
| Device-related thrombosis and embolism | $\leq 10^a$ | $\leq 10^a$ | 1.003 (0.412, 2.439) | 0.316 |
| **Thoracic complications** |  |  |  |  |
| Haemothorax | $\leq 10^a$ | $\leq 10^a$ | 0.997 (0.410, 2.426) | 0.995 |
| Pneumothorax | $\leq 10^a$ | 0 | — | 0.001** |

### Device-related complications

Device-related complications occurred at low rates in both groups and were observed in ≤10 patients in the LLPM group and in 20 patients (5.5%) in the TVPM group. No statistically significant difference was found (OR 0.49, 95% CI 0.23–1.07; p = 0.067). Device-related thrombosis and embolism occurred in ≤10 patients in both groups.

Other complications, including unspecified non-cardiac or systemic events, were reported in ≤10 patients in both groups, without significant difference (OR 1.01, 95% CI 0.42–2.46; p = 0.979).

Due to the limited number of events (≤10 in at least one group), several outcomes in this category did not allow for reliable estimation of odds or hazard ratios. These values are displayed as “≤10” in accordance with TriNetX data suppression policies.

## Discussion

In this large multicenter real-world study, we found that LLPM were associated with a significantly lower incidence of heart failure and atrial fibrillation compared to TVPM in patients implanted after TAVR, while all-cause mortality and complication rates remained similar between groups.

Our findings complement and expand upon those of Ueyama et al., who recently conducted a large Medicare-based analysis comparing outcomes of leadless and transvenous pacemakers following TAVR.^23^ Notably, their population included both single- and dual-chamber transvenous systems followed up to two years, introducing potential bias due to shorter follow-up duration and variations in atrioventricular synchrony. In contrast, our study focused exclusively on single-chamber ventricular pacing (VVI/VVIR), thereby removing confounding effects related to pacing mode or complications related to atrial lead. This methodological choice likely enhances the internal validity of our findings and isolates the impact of the pacing system itself, rather than pacing strategy.

All-cause mortality and procedural or device-related complication rates were similar between groups. When considering mortality, this is consistent with results previously reported by Ueyama et al. in post-TAVR patients and also in high-risk patients as described by Boveda et al. where no difference was reported in each group.^23,26^ Patients undergoing TAVR experience an early and high mortality rate with 18% of patients dying within the first year and with a yearly incidence of 15.5% mainly driven by non-cardiovascular death.^10^ In addition, post-TAVR pacemaker implantation is associated with an increased mortality.^12^ This induces a competing risk masking mid- or long-term device-related complications and subsequent mortality. Hence, we did not report any difference in terms of device-related complications between LLPMs and TVPMs. Our findings are consistent with those of Ueyama et al. who reported no difference when LLPM were compared to the subgroup of patients implanted with only VVI-TVPM and showed the difference in the overall cohort was mainly driven by dual-chamber pacemakers.^23^ This underlines the paradox that patients who may benefit the most from LLPMs, as reported for end-stage renal disease by Boveda et al., actually do not because of a shorter life expectancy.^26^ These findings have important clinical and economic implications, especially regarding the vulnerability of the post-TAVR population—elderly, frail, and often burdened with multiple comorbidities.^6,8,27^

These neutral results should not overshadow the potential benefits in morbidity outcomes. Previous nationwide analyses have demonstrated that permanent pacing after TAVR can negatively influence outcomes, especially when performed after balloon-expandable valves or in patients with limited baseline functional reserve.^11,12^ These findings support our hypothesis that pacing modality—and not merely the need for pacing—may significantly affect the long-term prognosis of post-TAVR patients.

The reduction in heart failure observed in the LLPM group may reflect the deleterious effects of conventional right ventricular apical pacing, which can promote ventricular dyssynchrony and worsen left ventricular function. ^21,22,26,28^ Leadless systems, typically deployed in the septal region of the right ventricle, may mitigate this risk and therefore reduce the occurrence of pacing induced cardiomyopathy.^29,30^ Moreover, the absence of leads reduces tricuspid valve interaction and the associated risk of regurgitation.^31–33^

The observed reduction in atrial fibrillation in the LLPM cohort also merits discussion. While atrial fibrillation is often pre-existing in the TAVR population, incident cases may reflect atrial remodeling induced by pacing-induced dyssynchrony. Right ventricular pacing has been linked to increased atrial pressures and mechanical dispersion, favoring the development of arrhythmias.^34,35^ The reduced atrial burden in LLPM recipients may thus reflect a more physiological hemodynamic profile possibly due to the same mechanisms as discussed above for heart failure reduction. However, we acknowledge the limitation of not having data on atrial arrhythmia burden or precise timing relative to implantation.

In a population for which quality of life, hospitalizations, and functional status are critical endpoints, reducing heart failure and atrial arrhythmias is highly meaningful. Mortality as an endpoint may be insensitive to these intermediate improvements, especially in elderly populations with competing risks.^20,21,23,36^

Regarding safety, both pacing strategies showed low and comparable rates of complications. Notably, pneumothorax was observed in the LLPM group—a finding likely explained by prior unsuccessful transvenous attempts or misclassification in procedural coding.^20,37^ This underscores a common limitation of claims-based datasets: the inability to establish procedural chronology or granularity in causality. Similarly, while we included endocarditis as an exploratory outcome, it remains difficult to attribute these events specifically to the device type in the absence of microbiological or echocardiographic data.

These findings open the door to several clinical and research perspectives. First, LLPM may be particularly appealing in subgroups at high risk for device-related complications, such as patients with chronic kidney disease, prior device infections, or limited vascular access.^25,38^ Our data support the hypothesis that in such populations, leadless pacing could improve clinical outcomes and reduce post-procedural morbidity. Second, the potential for LLPM to reduce pacing-induced cardiomyopathy—through both septal placement and minimized interference with tricuspid anatomy—should be further explored in dedicated studies.

Additionally, ongoing technological advances in leadless pacing, including the development of dual-chamber systems, algorithms for AV synchrony (e.g., MICRA AV, AVEIR DR)^39^, and even early iterations of leadless conduction system pacing (CSP) systems^40^, will likely expand the indications for leadless systems in broader patient populations. Meanwhile, cost-effectiveness analyses and health economic evaluations will be essential to guide adoption strategies, especially given the current cost differential between leadless and conventional systems.

Prospective randomized trials remain needed to confirm the safety and efficacy of LLPM in the post-TAVR population. Future studies should ideally integrate pacing burden, device programming details, echocardiographic parameters (e.g., ventricular function, valve regurgitation), and patient-reported outcomes. The inclusion of imaging-based endpoints could also clarify the impact of pacing site on chamber remodeling and function.

In conclusion, in a propensity-matched cohort of post-TAVR patients requiring permanent pacing, LLPMs were associated with a reduced risk of heart failure and atrial fibrillation, while maintaining a comparable safety and mortality profile to conventional VVI transvenous devices. This study contributes real-world evidence supporting the continued expansion of leadless technology in contemporary structural heart disease management.

## Acknowledgments

None

## Ethics statement

This study used de-identified electronic health records from the TriNetX Global Collaborative Network. In accordance with participating institutions’ policies, analyses of de-identified data were determined to be non–human subjects research; institutional review board approval and individual informed consent were not required.

## Data availability

Data were accessed under license via the TriNetX Global Collaborative Network. De-identified aggregate results supporting this study are available from the corresponding author on reasonable request; patient-level data cannot be shared publicly under data-use agreements.

## Supplemental Material

**Figure S1.**
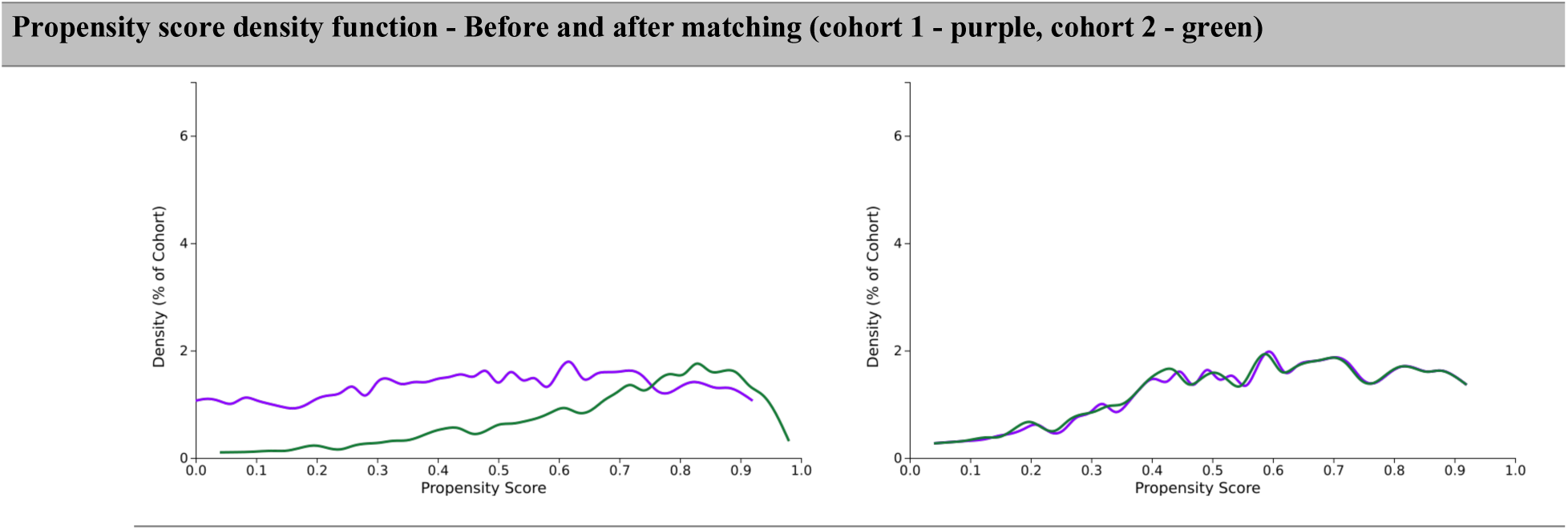
Propensity score density function before and after matching.

**Figure S2.**
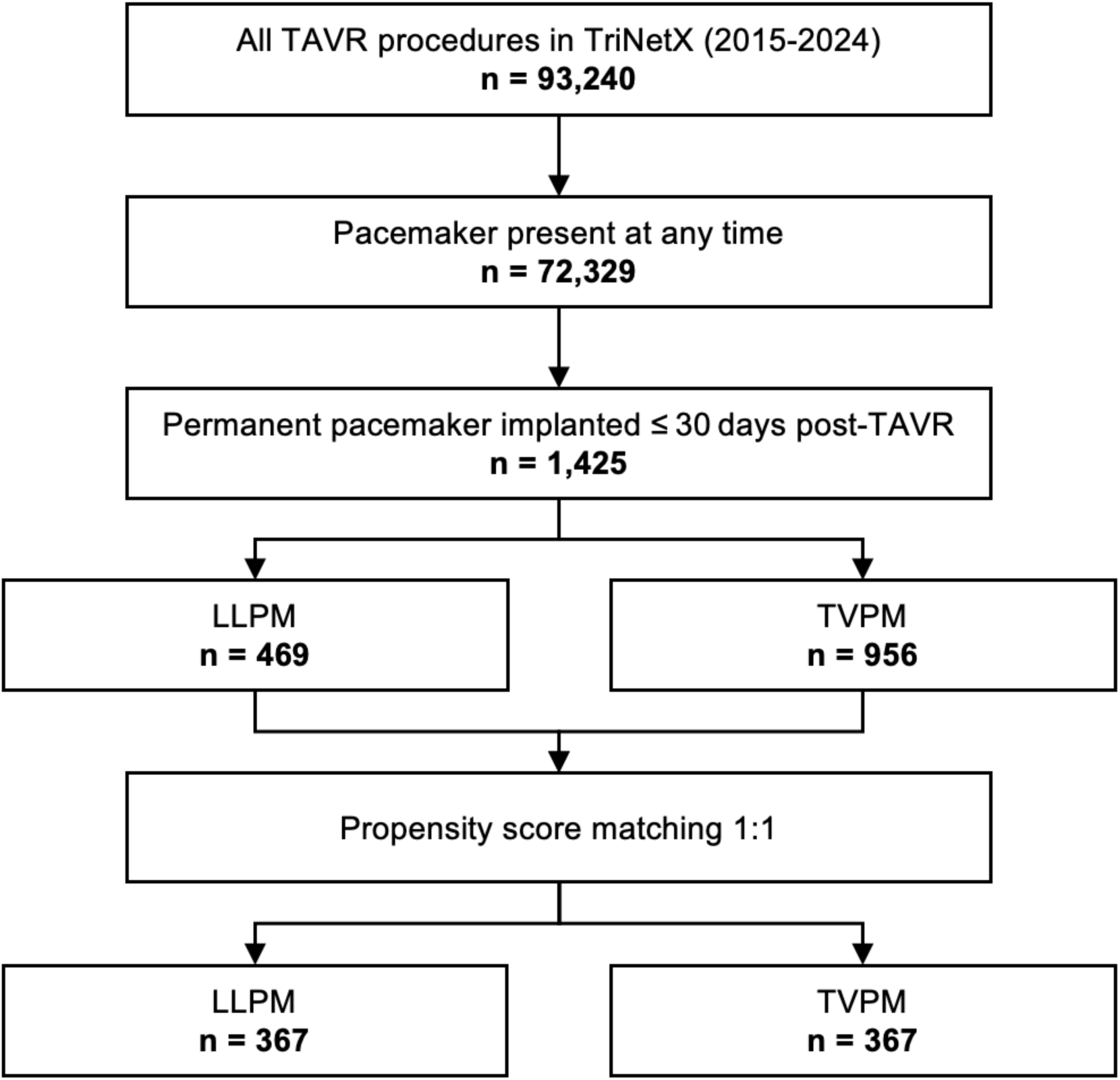
Patient selection flowchart.

Among all patients who underwent transcatheter aortic valve replacement (TAVR) in the TriNetX network between 2015 and 2024 (N = 93,240), 72,329 had a pacemaker present at any time (pre-existing or implanted > 30 days post-TAVR). The study cohort included 1,425 patients who underwent new permanent pacemaker implantation within 30 days after TAVR, divided into leadless (LLPM, n = 469) and transvenous single-chamber (TVPM, n = 956) groups. One-to-one propensity score matching yielded two matched cohorts of 367 patients each.

## Supplemental Methods

### Appendix A Text Representation of the Cohorts Definition

This section lists all terms used in the definitions of the two cohorts.

#### Query Criteria for Cohort 1 (query name: LLPM)

Patients must have:

Age (Age) (at least 50 years (most recent occurrence)).

All the following must be satisfied:

LLPM: The terms in this group occurred between Jan 1, 2015 and Dec 31, 2024

Patients must have:

any of the following:

Transcatheter insertion or replacement of permanent leadless pacemaker, right ventricular, including imaging guidance (eg, fluoroscopy, venous ultrasound, ventriculography, femoral venography) and device evaluation (eg, interrogation or programming), when performed (UMLS:CPT:33274); or

Transcatheter insertion or replacement of permanent leadless pacemaker, ventricular (deprecated 2020) (UMLS:CPT:0387T); or

Insertion of Intracardiac Pacemaker into Right Ventricle, Percutaneous Approach (UMLS:ICD10PCS:02HK3NZ).

TAVI: The first instance of TAVI occurred within 1 month on or before the first instance of LLPM

Patients must have:

any of the following:

Transcatheter aortic valve replacement (TAVR/TAVI) with prosthetic valve (UMLS:CPT:1021150); or

Replacement of Aortic Valve with Autologous Tissue Substitute, Transapical, Percutaneous Approach (UMLS:ICD10PCS:02RF37H); or

Replacement of Aortic Valve with Autologous Tissue Substitute, Percutaneous Approach (UMLS:ICD10PCS:02RF37Z); or

Replacement of Aortic Valve with Zooplastic Tissue, Transapical, Percutaneous Approach (UMLS:ICD10PCS:02RF38H); or

Replacement of Aortic Valve with Zooplastic Tissue, Percutaneous Approach (UMLS:ICD10PCS:02RF38Z); or

Replacement of Aortic Valve with Synthetic Substitute, Transapical, Percutaneous Approach (UMLS:ICD10PCS:02RF3JH); or

Replacement of Aortic Valve with Synthetic Substitute, Percutaneous Approach (UMLS:ICD10PCS:02RF3JZ); or

Replacement of Aortic Valve with Nonautologous Tissue Substitute, Transapical, Percutaneous Approach (UMLS:ICD10PCS:02RF3KH); or

Replacement of Aortic Valve with Nonautologous Tissue Substitute, Percutaneous Approach (UMLS:ICD10PCS:02RF3KZ).

Patients cannot have:

any of the following:

Presence of cardiac pacemaker (UMLS:ICD10CM:Z95.0); or

Presence of automatic (implantable) cardiac defibrillator (UMLS:ICD10CM:Z95.810).

#### Query Criteria for Cohort 2 (query name: TVPM)

Patients must have:

Age (Age) (at least 50 years (most recent occurrence)).

All the following must be satisfied:

Any of the following must be satisfied:

TVPM: The terms in this group occurred between Jan 1, 2015 and Dec 31, 2024

Patients must have:

any of the following:

Pacemaker, single chamber, rate-responsive (implantable) (UMLS:HCPCS:C1786);

or

Pacemaker, single chamber, non rate-responsive (implantable) (UMLS:HCPCS:C2620).

TAVI: The first instance of TAVI occurred within 1 month on or before the first instance of TVPM

Patients must have:

any of the following:

Replacement of Aortic Valve with Autologous Tissue Substitute, Transapical, Percutaneous Approach (UMLS:ICD10PCS:02RF37H); or

Replacement of Aortic Valve with Autologous Tissue Substitute, Percutaneous Approach (UMLS:ICD10PCS:02RF37Z); or

Replacement of Aortic Valve with Zooplastic Tissue, Transapical, Percutaneous Approach (UMLS:ICD10PCS:02RF38H); or

Replacement of Aortic Valve with Zooplastic Tissue, Percutaneous Approach (UMLS:ICD10PCS:02RF38Z); or

Replacement of Aortic Valve with Synthetic Substitute, Transapical, Percutaneous Approach (UMLS:ICD10PCS:02RF3JH); or

Replacement of Aortic Valve with Synthetic Substitute, Percutaneous Approach (UMLS:ICD10PCS:02RF3JZ); or

Replacement of Aortic Valve with Nonautologous Tissue Substitute, Transapical, Percutaneous Approach (UMLS:ICD10PCS:02RF3KH); or

Replacement of Aortic Valve with Nonautologous Tissue Substitute, Percutaneous Approach (UMLS:ICD10PCS:02RF3KZ).

Patients cannot have:

any of the following:

Presence of cardiac pacemaker (UMLS:ICD10CM:Z95.0); or

Presence of automatic (implantable) cardiac defibrillator

(UMLS:ICD10CM:Z95.810).

VVI 2: The terms in this group occurred between Jan 1, 2015 and Dec 31, 2024

Patients must have:

all of the following:

Insertion of Pacemaker, Single Chamber into Chest Subcutaneous Tissue and Fascia, Open Approach (UMLS:ICD10PCS:0JH604Z); and

Insertion of Pacemaker Lead into Right Ventricle, Percutaneous Approach (UMLS:ICD10PCS:02HK3JZ).

TAVI: The first instance of TAVI occurred within 1 month on or before the first instance of VVI 2

Patients must have:

any of the following:

Replacement of Aortic Valve with Autologous Tissue Substitute, Transapical, Percutaneous Approach (UMLS:ICD10PCS:02RF37H); or

Replacement of Aortic Valve with Autologous Tissue Substitute, Percutaneous Approach (UMLS:ICD10PCS:02RF37Z); or

Replacement of Aortic Valve with Zooplastic Tissue, Transapical, Percutaneous Approach (UMLS:ICD10PCS:02RF38H); or

Replacement of Aortic Valve with Zooplastic Tissue, Percutaneous Approach (UMLS:ICD10PCS:02RF38Z); or

Replacement of Aortic Valve with Synthetic Substitute, Transapical, Percutaneous Approach (UMLS:ICD10PCS:02RF3JH); or

Replacement of Aortic Valve with Synthetic Substitute, Percutaneous Approach (UMLS:ICD10PCS:02RF3JZ); or

Replacement of Aortic Valve with Nonautologous Tissue Substitute, Transapical, Percutaneous Approach (UMLS:ICD10PCS:02RF3KH); or

Replacement of Aortic Valve with Nonautologous Tissue Substitute, Percutaneous Approach (UMLS:ICD10PCS:02RF3KZ).

Patients cannot have:

any of the following:

Presence of cardiac pacemaker (UMLS:ICD10CM:Z95.0); or

Presence of automatic (implantable) cardiac defibrillator (UMLS:ICD10CM:Z95.810).

VVI 3: The terms in this group occurred between Jan 1, 2015 and Dec 31, 2024

Patients must have:

all of the following:

Insertion of Pacemaker, Single Chamber Rate Responsive into Chest Subcutaneous Tissue and Fascia, Open Approach (UMLS:ICD10PCS:0JH605Z); and

Insertion of Pacemaker Lead into Right Ventricle, Percutaneous Approach (UMLS:ICD10PCS:02HK3JZ).

TAVI: The first instance of TAVI occurred within 1 month on or before the first instance of VVI 3

Patients must have:

any of the following:

Replacement of Aortic Valve with Autologous Tissue Substitute, Transapical, Percutaneous Approach (UMLS:ICD10PCS:02RF37H); or

Replacement of Aortic Valve with Autologous Tissue Substitute, Percutaneous Approach (UMLS:ICD10PCS:02RF37Z); or

Replacement of Aortic Valve with Zooplastic Tissue, Transapical, Percutaneous Approach (UMLS:ICD10PCS:02RF38H); or

Replacement of Aortic Valve with Zooplastic Tissue, Percutaneous Approach (UMLS:ICD10PCS:02RF38Z); or

Replacement of Aortic Valve with Synthetic Substitute, Transapical, Percutaneous Approach (UMLS:ICD10PCS:02RF3JH); or

Replacement of Aortic Valve with Synthetic Substitute, Percutaneous Approach (UMLS:ICD10PCS:02RF3JZ); or

Replacement of Aortic Valve with Nonautologous Tissue Substitute, Transapical, Percutaneous Approach (UMLS:ICD10PCS:02RF3KH); or

Replacement of Aortic Valve with Nonautologous Tissue Substitute, Percutaneous Approach (UMLS:ICD10PCS:02RF3KZ).

Patients cannot have:

any of the following:

Presence of cardiac pacemaker (UMLS:ICD10CM:Z95.0); or

Presence of automatic (implantable) cardiac defibrillator (UMLS:ICD10CM:Z95.810).

### Appendix B Text Representation of the Analysis Setup

This section contains the Index Event definition for each cohort.

The index event for Cohort 1 (query name: LLPM) is defined as the following:

All the following must be satisfied:

LPM: The terms in this group occurred between Jan 1, 2015 and Dec 31, 2024

Patients must have:

any of the following:

Transcatheter insertion or replacement of permanent leadless pacemaker, right ventricular, including imaging guidance (eg, fluoroscopy, venous ultrasound, ventriculography, femoral venography) and device evaluation (eg, interrogation or programming), when performed (UMLS:CPT:33274); or

Transcatheter insertion or replacement of permanent leadless pacemaker, ventricular (deprecated 2020) (UMLS:CPT:0387T); or

Insertion of Intracardiac Pacemaker into Right Ventricle, Percutaneous Approach (UMLS:ICD10PCS:02HK3NZ).

TAVI: The first instance of TAVI occurred within 1 month on or before the first instance of LPM

Patients must have:

any of the following:

Transcatheter aortic valve replacement (TAVR/TAVI) with prosthetic valve (UMLS:CPT:1021150); or

Replacement of Aortic Valve with Autologous Tissue Substitute, Transapical, Percutaneous Approach (UMLS:ICD10PCS:02RF37H); or

Replacement of Aortic Valve with Autologous Tissue Substitute, Percutaneous Approach (UMLS:ICD10PCS:02RF37Z); or

Replacement of Aortic Valve with Zooplastic Tissue, Transapical, Percutaneous Approach (UMLS:ICD10PCS:02RF38H); or

Replacement of Aortic Valve with Zooplastic Tissue, Percutaneous Approach (UMLS:ICD10PCS:02RF38Z); or

Replacement of Aortic Valve with Synthetic Substitute, Transapical, Percutaneous Approach (UMLS:ICD10PCS:02RF3JH); or

Replacement of Aortic Valve with Synthetic Substitute, Percutaneous Approach (UMLS:ICD10PCS:02RF3JZ); or

Replacement of Aortic Valve with Nonautologous Tissue Substitute, Transapical, Percutaneous Approach (UMLS:ICD10PCS:02RF3KH); or

Replacement of Aortic Valve with Nonautologous Tissue Substitute, Percutaneous Approach (UMLS:ICD10PCS:02RF3KZ).

Patients cannot have: any of the following:

Presence of cardiac pacemaker (UMLS:ICD10CM:Z95.0); or

Presence of automatic (implantable) cardiac defibrillator (UMLS:ICD10CM:Z95.810).

The index event for Cohort 2 (query name: TVPM) is defined as the following: All the following must be satisfied:

Any of the following must be satisfied:

TVPM: The terms in this group occurred between Jan 1, 2015 and Dec 31, 2024

Patients must have:

any of the following:

Pacemaker, single chamber, rate-responsive (implantable) (UMLS:HCPCS:C1786); or

Pacemaker, single chamber, non rate-responsive (implantable) (UMLS:HCPCS:C2620).

TAVI: The first instance of TAVI occurred within 1 month on or before the first instance of TVPM

Patients must have:

any of the following:

Replacement of Aortic Valve with Autologous Tissue Substitute, Transapical, Percutaneous Approach (UMLS:ICD10PCS:02RF37H); or

Replacement of Aortic Valve with Autologous Tissue Substitute, Percutaneous Approach (UMLS:ICD10PCS:02RF37Z); or

Replacement of Aortic Valve with Zooplastic Tissue, Transapical, Percutaneous Approach (UMLS:ICD10PCS:02RF38H); or

Replacement of Aortic Valve with Zooplastic Tissue, Percutaneous Approach (UMLS:ICD10PCS:02RF38Z); or

Replacement of Aortic Valve with Synthetic Substitute, Transapical, Percutaneous Approach (UMLS:ICD10PCS:02RF3JH); or

Replacement of Aortic Valve with Synthetic Substitute, Percutaneous Approach (UMLS:ICD10PCS:02RF3JZ); or

Replacement of Aortic Valve with Nonautologous Tissue Substitute, Transapical, Percutaneous Approach (UMLS:ICD10PCS:02RF3KH); or

Replacement of Aortic Valve with Nonautologous Tissue Substitute, Percutaneous Approach (UMLS:ICD10PCS:02RF3KZ).

Patients cannot have:

any of the following:

Presence of cardiac pacemaker (UMLS:ICD10CM:Z95.0); or

Presence of automatic (implantable) cardiac defibrillator (UMLS:ICD10CM:Z95.810).

VVI 2: The terms in this group occurred between Jan 1, 2015 and Dec 31, 2024

Patients must have:

all of the following:

Insertion of Pacemaker, Single Chamber into Chest Subcutaneous Tissue and Fascia, Open Approach (UMLS:ICD10PCS:0JH604Z); and

Insertion of Pacemaker Lead into Right Ventricle, Percutaneous Approach (UMLS:ICD10PCS:02HK3JZ).

TAVI: The first instance of TAVI occurred within 1 month on or before the first instance of VVI 2

Patients must have:

any of the following:

Replacement of Aortic Valve with Autologous Tissue Substitute, Transapical, Percutaneous Approach (UMLS:ICD10PCS:02RF37H); or

Replacement of Aortic Valve with Autologous Tissue Substitute, Percutaneous Approach (UMLS:ICD10PCS:02RF37Z); or

Replacement of Aortic Valve with Zooplastic Tissue, Transapical, Percutaneous Approach (UMLS:ICD10PCS:02RF38H); or

Replacement of Aortic Valve with Zooplastic Tissue, Percutaneous Approach (UMLS:ICD10PCS:02RF38Z); or

Replacement of Aortic Valve with Synthetic Substitute, Transapical, Percutaneous Approach (UMLS:ICD10PCS:02RF3JH); or

Replacement of Aortic Valve with Synthetic Substitute, Percutaneous Approach (UMLS:ICD10PCS:02RF3JZ); or

Replacement of Aortic Valve with Nonautologous Tissue Substitute, Transapical, Percutaneous Approach (UMLS:ICD10PCS:02RF3KH); or

Replacement of Aortic Valve with Nonautologous Tissue Substitute, Percutaneous Approach (UMLS:ICD10PCS:02RF3KZ).

Patients cannot have:

any of the following:

Presence of cardiac pacemaker (UMLS:ICD10CM:Z95.0); or

Presence of automatic (implantable) cardiac defibrillator

(UMLS:ICD10CM:Z95.810).

VVI 3: The terms in this group occurred between Jan 1, 2015 and Dec 31, 2024

Patients must have:

all of the following:

Insertion of Pacemaker, Single Chamber Rate Responsive into Chest Subcutaneous Tissue and Fascia, Open Approach (UMLS:ICD10PCS:0JH605Z); and

Insertion of Pacemaker Lead into Right Ventricle, Percutaneous Approach (UMLS:ICD10PCS:02HK3JZ).

TAVI: The first instance of TAVI occurred within 1 month on or before the first instance of VVI 3

Patients must have:

any of the following:

Replacement of Aortic Valve with Autologous Tissue Substitute, Transapical, Percutaneous Approach (UMLS:ICD10PCS:02RF37H); or

Replacement of Aortic Valve with Autologous Tissue Substitute, Percutaneous Approach (UMLS:ICD10PCS:02RF37Z); or

Replacement of Aortic Valve with Zooplastic Tissue, Transapical, Percutaneous Approach (UMLS:ICD10PCS:02RF38H); or

Replacement of Aortic Valve with Zooplastic Tissue, Percutaneous Approach (UMLS:ICD10PCS:02RF38Z); or

Replacement of Aortic Valve with Synthetic Substitute, Transapical, Percutaneous Approach (UMLS:ICD10PCS:02RF3JH); or

Replacement of Aortic Valve with Synthetic Substitute, Percutaneous Approach (UMLS:ICD10PCS:02RF3JZ); or

Replacement of Aortic Valve with Nonautologous Tissue Substitute, Transapical, Percutaneous Approach (UMLS:ICD10PCS:02RF3KH); or

Replacement of Aortic Valve with Nonautologous Tissue Substitute, Percutaneous Approach (UMLS:ICD10PCS:02RF3KZ).

Patients cannot have:

any of the following:

Presence of cardiac pacemaker (UMLS:ICD10CM:Z95.0); or

Presence of automatic (implantable) cardiac defibrillator

(UMLS:ICD10CM:Z95.810).

### Appendix C Text Representation of the Outcomes Definition

This analysis includes the following outcomes:

Death

Patients must have:

any of the following:

Ill-defined and unknown cause of mortality (UMLS:ICD10CM:R99); or

Deceased (Deceased).

Puncture site complication

Patients must have:

any of the following:

Arteriovenous fistula, acquired (UMLS:ICD10CM:I77.0); or

Aneurysm of iliac artery (UMLS:ICD10CM:I72.3); or

Aneurysm of other specified arteries (UMLS:ICD10CM:I72.8); or

Aneurysm of unspecified site (UMLS:ICD10CM:I72.9); or

Aneurysm of artery of lower extremity (UMLS:ICD10CM:I72.4).

Heart failure

Patients must have:

Heart failure (UMLS:ICD10CM:I50) (principalIndicator: Primary Priority).

Cardiac effusion or perforation

Patients must have:

any of the following:

Hemopericardium, not elsewhere classified (UMLS:ICD10CM:I31.2); or

Cardiac tamponade (UMLS:ICD10CM:I31.4); or

Pericardial effusion (noninflammatory) (UMLS:ICD10CM:I31.3).

Venous embolism and thrombosis

Patients must have:

any of the following:

Other venous embolism and thrombosis (UMLS:ICD10CM:I82); or

Pulmonary embolism (UMLS:ICD10CM:I26).

Device related thrombosis and embolism

Patients must have:

any of the following:

Thrombosis due to cardiac prosthetic devices, implants and grafts (UMLS:ICD10CM:T82.867); or

Embolism due to cardiac prosthetic devices, implants and grafts (UMLS:ICD10CM:T82.817).

Atrial fibrillation

Patients must have:

Atrial fibrillation and flutter (UMLS:ICD10CM:I48) (principalIndicator: Primary Priority).

Hemothorax

Patients must have:

any of the following:

Postprocedural pneumothorax and air leak (UMLS:ICD10CM:J95.81); or

Hemothorax (UMLS:ICD10CM:J94.2); or

Intraoperative hemorrhage and hematoma of a respiratory system organ or structure complicating other procedure (UMLS:ICD10CM:J95.62); or

Accidental puncture and laceration of a respiratory system organ or structure during other procedure (UMLS:ICD10CM:J95.72); or

Postprocedural pneumothorax and air leak (UMLS:ICD10CM:J95.81).

Pneumothorax

Patients must have:

any of the following:

Postprocedural pneumothorax (UMLS:ICD10CM:J95.811); or

Postprocedural air leak (UMLS:ICD10CM:J95.812).

Pericarditis

Patients must have:

any of the following:

Acute pericarditis, unspecified (UMLS:ICD10CM:I30.9); or

Disease of pericardium, unspecified (UMLS:ICD10CM:I31.9).

Intraoperative cardiac arrest Patients must have:

any of the following:

Intraoperative cardiac arrest during cardiac surgery (UMLS:ICD10CM:I97.710); or

Postprocedural cardiac arrest following cardiac surgery (UMLS:ICD10CM:I97.120).

Vascular complication

Patients must have:

any of the following:

Intraoperative hemorrhage and hematoma of a circulatory system organ or structure complicating other circulatory system procedure (UMLS:ICD10CM:I97.418); or

Postprocedural hemorrhage of a circulatory system organ or structure following other circulatory system procedure (UMLS:ICD10CM:I97.618).

Endocarditis

Patients must have:

any of the following:

Acute and subacute infective endocarditis (UMLS:ICD10CM:I33.0); or

Endocarditis, valve unspecified (UMLS:ICD10CM:I38); or

Acute and subacute endocarditis, unspecified (UMLS:ICD10CM:I33.9); or

Endocarditis and heart valve disorders in diseases classified elsewhere (UMLS:ICD10CM:I39); or

Candidal endocarditis (UMLS:ICD10CM:B37.6); or

Meningococcal endocarditis (UMLS:ICD10CM:A39.51); or

Gonococcal heart infection (UMLS:ICD10CM:A54.83); or

Infection and inflammatory reaction due to cardiac valve prosthesis (UMLS:ICD10CM:T82.6); or

Infection and inflammatory reaction due to cardiac valve prosthesis, subsequent encounter (UMLS:ICD10CM:T82.6XXD); or

Infection and inflammatory reaction due to cardiac valve prosthesis, initial encounter (UMLS:ICD10CM:T82.6XXA).

Other complications

Patients must have:

any of the following:

Postprocedural hematoma of a circulatory system organ or structure following other circulatory system procedure (UMLS:ICD10CM:I97.638); or

Postprocedural hemorrhage of a circulatory system organ or structure following other circulatory system procedure (UMLS:ICD10CM:I97.618); or

Intraoperative cardiac arrest during cardiac surgery (UMLS:ICD10CM:I97.710); or

Postprocedural cardiac arrest following cardiac surgery (UMLS:ICD10CM:I97.120); or

Acute pericarditis, unspecified (UMLS:ICD10CM:I30.9); or

Disease of pericardium, unspecified (UMLS:ICD10CM:I31.9); or

Intraoperative hemorrhage and hematoma of a circulatory system organ or structure complicating other circulatory system procedure (UMLS:ICD10CM:I97.418); or

Postprocedural hemorrhage of a circulatory system organ or structure following other circulatory system procedure (UMLS:ICD10CM:I97.618); or

Intraoperative hemorrhage and hematoma of a respiratory system organ or structure complicating other procedure (UMLS:ICD10CM:J95.62); or

Postprocedural hemorrhage of a respiratory system organ or structure following other procedure (UMLS:ICD10CM:J95.831); or

Accidental puncture and laceration of a respiratory system organ or structure during other procedure (UMLS:ICD10CM:J95.72); or

Hemothorax (UMLS:ICD10CM:J94.2); or

Postprocedural pneumothorax (UMLS:ICD10CM:J95.811); or

Postprocedural air leak (UMLS:ICD10CM:J95.812).

Device related complications

Patients must have:

any of the following:

Breakdown (mechanical) of cardiac electronic device (UMLS:ICD10CM:T82.11); or

Displacement of cardiac electronic device (UMLS:ICD10CM:T82.12); or

Other mechanical complication of cardiac electronic device (UMLS:ICD10CM:T82.19); or

Infection and inflammatory reaction due to other cardiac and vascular devices, implants and grafts (UMLS:ICD10CM:T82.7); or

Hemorrhage due to cardiac prosthetic devices, implants and grafts (UMLS:ICD10CM:T82.837); or

Pain due to cardiac prosthetic devices, implants and grafts (UMLS:ICD10CM:T82.847); or

Stenosis of other cardiac prosthetic devices, implants and grafts (UMLS:ICD10CM:T82.857); or

Other specified complication of cardiac prosthetic devices, implants and grafts (UMLS:ICD10CM:T82.897).

Post procedural hematoma

Patients must have:

Postprocedural hematoma of a circulatory system organ or structure following other circulatory system procedure (UMLS:ICD10CM:I97.638).

Post procedural hemorrage

Patients must have:

Postprocedural hemorrhage of a circulatory system organ or structure following other circulatory system procedure (UMLS:ICD10CM:I97.618).

Overall complication (except HF&IE)

Patients must have:

any of the following:

Other venous embolism and thrombosis (UMLS:ICD10CM:I82); or

Pulmonary embolism (UMLS:ICD10CM:I26); or

Thrombosis due to cardiac prosthetic devices, implants and grafts (UMLS:ICD10CM:T82.867); or

Embolism due to cardiac prosthetic devices, implants and grafts (UMLS:ICD10CM:T82.817); or

Accidental puncture and laceration of a circulatory system organ or structure during a circulatory system procedure (UMLS:ICD10CM:I97.51); or

Hemopericardium, not elsewhere classified (UMLS:ICD10CM:I31.2); or

Pericardial effusion (noninflammatory) (UMLS:ICD10CM:I31.3); or

Cardiac tamponade (UMLS:ICD10CM:I31.4); or

Arteriovenous fistula, acquired (UMLS:ICD10CM:I77.0); or

Aneurysm of artery of lower extremity (UMLS:ICD10CM:I72.4); or

Breakdown (mechanical) of cardiac electronic device (UMLS:ICD10CM:T82.11); or

Displacement of cardiac electronic device (UMLS:ICD10CM:T82.12); or

Other mechanical complication of cardiac electronic device (UMLS:ICD10CM:T82.19); or

Infection and inflammatory reaction due to other cardiac and vascular devices, implants and grafts (UMLS:ICD10CM:T82.7); or

Hemorrhage due to cardiac prosthetic devices, implants and grafts (UMLS:ICD10CM:T82.837); or

Pain due to cardiac prosthetic devices, implants and grafts (UMLS:ICD10CM:T82.847); or

Stenosis of other cardiac prosthetic devices, implants and grafts (UMLS:ICD10CM:T82.857); or

Other specified complication of cardiac prosthetic devices, implants and grafts (UMLS:ICD10CM:T82.897); or

Postprocedural hematoma of a circulatory system organ or structure following other circulatory system procedure (UMLS:ICD10CM:I97.638); or

Postprocedural hemorrhage of a circulatory system organ or structure following other circulatory system procedure (UMLS:ICD10CM:I97.618); or

Intraoperative cardiac arrest during cardiac surgery (UMLS:ICD10CM:I97.710); or

Postprocedural cardiac arrest following cardiac surgery (UMLS:ICD10CM:I97.120); or

Acute pericarditis, unspecified (UMLS:ICD10CM:I30.9); or

Disease of pericardium, unspecified (UMLS:ICD10CM:I31.9); or

Intraoperative hemorrhage and hematoma of a circulatory system organ or structure complicating other circulatory system procedure (UMLS:ICD10CM:I97.418); or

Intraoperative hemorrhage and hematoma of a respiratory system organ or structure complicating other procedure (UMLS:ICD10CM:J95.62); or

Postprocedural hemorrhage of a respiratory system organ or structure following other procedure (UMLS:ICD10CM:J95.831); or

Accidental puncture and laceration of a respiratory system organ or structure during other procedure (UMLS:ICD10CM:J95.72); or

Hemothorax (UMLS:ICD10CM:J94.2); or

Postprocedural pneumothorax (UMLS:ICD10CM:J95.811); or

Postprocedural air leak (UMLS:ICD10CM:J95.812).

Tricuspid insufficiency or management

Patients must have:

any of the following:

Nonrheumatic tricuspid valve disorders (UMLS:ICD10CM:I36); or

Tricuspid Valve Repair (UMLS:CPT:1035630); or

Tricuspid Valve Implantation/Replacement (UMLS:CPT:1036591); or

Tricuspid annuloplasty using ring (UMLS:SNOMED:232782007); or

Valvuloplasty, tricuspid valve; with ring insertion (UMLS:CPT:33464); or

Valvuloplasty, tricuspid valve; without ring insertion (UMLS:CPT:33463); or

Repair Tricuspid Valve, Open Approach (UMLS:ICD10PCS:02QJ0ZZ); or

Surgical Procedures on the Tricuspid Valve (UMLS:CPT:1006167); or

Valvuloplasty, tricuspid valve (UMLS:CPT:1006169); or

Replacement of Tricuspid Valve with Autologous Tissue Substitute, Open Approach (UMLS:ICD10PCS:02RJ07Z); or

Replacement of Tricuspid Valve with Zooplastic Tissue, Percutaneous Endoscopic Approach (UMLS:ICD10PCS:02RJ48Z); or

Replacement of Tricuspid Valve with Autologous Tissue Substitute, Percutaneous Endoscopic Approach (UMLS:ICD10PCS:02RJ47Z); or

Replacement of Tricuspid Valve with Nonautologous Tissue Substitute, Percutaneous Endoscopic Approach (UMLS:ICD10PCS:02RJ4KZ); or

Replacement of Tricuspid Valve with Synthetic Substitute, Percutaneous Endoscopic Approach (UMLS:ICD10PCS:02RJ4JZ).

Tricuspid valve insuficiency

Patients must have:

Nonrheumatic tricuspid valve disorders (UMLS:ICD10CM:I36).

## References

1. Leon MB, Smith CR, Mack M, et al. Transcatheter aortic-valve implantation for aortic stenosis in patients who cannot undergo surgery. N Engl J Med. 2010;363(17):1597–1607. doi:10.1056/NEJMoa1008232

2. Leon MB, Smith CR, Mack MJ, et al. Transcatheter or Surgical Aortic-Valve Replacement in Intermediate-Risk Patients. N Engl J Med. 2016;374(17):1609–1620. doi:10.1056/NEJMoa1514616

3. Mack MJ, Leon MB, Thourani VH, et al. Transcatheter Aortic-Valve Replacement in Low-Risk Patients at Five Years. N Engl J Med. 2023;389(21):1949–1960. doi:10.1056/NEJMoa2307447

4. Cribier A, Eltchaninoff H, Bash A, et al. Percutaneous transcatheter implantation of an aortic valve prosthesis for calcific aortic stenosis: first human case description. Circulation. 2002;106(24):3006–3008. doi:10.1161/01.cir.0000047200.36165.b8

5. Sammour Y, Krishnaswamy A, Kumar A, et al. Incidence, Predictors, and Implications of Permanent Pacemaker Requirement After Transcatheter Aortic Valve Replacement. JACC Cardiovasc Interv. 2021;14(2):115–134. doi:10.1016/j.jcin.2020.09.063

6. Jelisejevas J, Regoli F, Hofer D, et al. Leadless Pacemaker Implantation, Focusing on Patients With Conduction System Disorders Post-Transcatheter Aortic Valve Replacement: A Retrospective Analysis. CJC Open. 2024;6(2Part A):96–103. doi:10.1016/j.cjco.2023.10.009

7. Nuche J, Ellenbogen KA, Mittal S, et al. Conduction Disturbances After Transcatheter Aortic Valve Replacement. JACC Cardiovasc Interv. 2024;17(22):2575–2595. doi:10.1016/j.jcin.2024.07.032

8. Lilly SM, Deshmukh AJ, Epstein AE, et al. 2020 ACC Expert Consensus Decision Pathway on Management of Conduction Disturbances in Patients Undergoing Transcatheter Aortic Valve Replacement. J Am Coll Cardiol. 2020;76(20):2391–2411. doi:10.1016/j.jacc.2020.08.050

9. Nuis RJ, Van Den Dorpel M, Adrichem R, Daemen J, Van Mieghem N. Conduction Abnormalities after Transcatheter Aortic Valve Implantation: Incidence, Impact and Management Using CT Data Interpretation. Interv Cardiol Rev Res Resour. 2024;19:e12. doi:10.15420/icr.2024.11

10. Lantelme P, Lacour T, Bisson A, et al. Futility Risk Model for Predicting Outcome After Transcatheter Aortic Valve Implantation. Am J Cardiol. 2020;130:100–107. doi:10.1016/j.amjcard.2020.05.043

11. Bisson A, Bodin A, Herbert J, et al. Pacemaker Implantation After Balloon- or Self-Expandable Transcatheter Aortic Valve Replacement in Patients With Aortic Stenosis. J Am Heart Assoc. 2020;9(9):e015896. doi:10.1161/JAHA.120.015896

12. Clementy N, Bisson A, Bodin A, et al. Outcomes associated with pacemaker implantation following transcatheter aortic valve replacement: A nationwide cohort study. Heart Rhythm. 2021;18(12):2027–2032. doi:10.1016/j.hrthm.2021.06.1175

13. Cantillon DJ, Derek V. Exner, et al. Complications and Health Care Costs Associated With Transvenous Cardiac Pacemakers in a Nationwide Assessment. JACC Clin Electrophysiol. 2017;3(11):1296–1305. doi:10.1016/j.jacep.2017.05.007

14. Cantillon DJ, Srinivas R. Dukkipati, et al. Comparative study of acute and mid-term complications with leadless and transvenous cardiac pacemakers. Heart Rhythm. 2018;15(7):1023–1030. doi:10.1016/j.hrthm.2018.04.022

15. Udo EO, Zuithoff NPA, Hemel NM van, et al. Incidence and predictors of short- and long-term complications in pacemaker therapy: The FOLLOWPACE study. Heart Rhythm. 2012;9(5):728–735. doi:10.1016/j.hrthm.2011.12.014

16. Duray GZ, Ritter P, El-Chami M, et al. Long-term performance of a transcatheter pacing system: 12-Month results from the Micra Transcatheter Pacing Study. Heart Rhythm. 2017;14(5):702–709. doi:10.1016/j.hrthm.2017.01.035

17. El-Chami MF, Soejima K, Piccini JP, et al. Incidence and outcomes of systemic infections in patients with leadless pacemakers: Data from the Micra IDE study. Pacing Clin Electrophysiol PACE. 2019;42(8):1105–1110. doi:10.1111/pace.13752

18. Mikhael F. El-Chami, El-Chami MF, Jens Brock Johansen, et al. Leadless pacemaker implant in patients with pre-existing infections: Results from the Micra postapproval registry. J Cardiovasc Electrophysiol. 2019;30(4):569–574. doi:10.1111/jce.13851

19. El-Chami MF, Al-Samadi F, Clementy N, et al. Updated performance of the Micra transcatheter pacemaker in the real-world setting: A comparison to the investigational study and a transvenous historical control. Heart Rhythm. 2018;15(12):1800–1807. doi:10.1016/j.hrthm.2018.08.005

20. El-Chami MF, Bockstedt L, Longacre C, et al. Leadless vs. transvenous single-chamber ventricular pacing in the Micra CED study: 2-year follow-up. Eur Heart J. 2022;43(12):1207–1215. doi:10.1093/eurheartj/ehab767

21. Crossley GH, Piccini JP, Longacre C, Higuera L, Stromberg K, El-Chami MF. Leadless versus transvenous single-chamber ventricular pacemakers: 3 year follow-up of the Micra CED study. J Cardiovasc Electrophysiol. 2023;34(4):1015–1023. doi:10.1111/jce.15863

22. Reynolds D, Duray GZ, Omar R, et al. A Leadless Intracardiac Transcatheter Pacing System. N Engl J Med. 2016;374(6):533–541. doi:10.1056/NEJMoa1511643

23. Ueyama HA, Miyamoto Y, Hashimoto K, et al. Comparison of Patient Outcomes Between Leadless vs Transvenous Pacemakers Following Transcatheter Aortic Valve Replacement. JACC Cardiovasc Interv. 2024;17(15):1779–1791. doi:10.1016/j.jcin.2024.05.030

24. Piccini JP, El-Chami M, Wherry K, et al. Contemporaneous Comparison of Outcomes Among Patients Implanted With a Leadless vs Transvenous Single-Chamber Ventricular Pacemaker. JAMA Cardiol. 2021;6(10):1187–1195. doi:10.1001/jamacardio.2021.2621

25. El-Chami MF, Clementy N, Garweg C, et al. Leadless Pacemaker Implantation in Hemodialysis Patients: Experience With the Micra Transcatheter Pacemaker. JACC Clin Electrophysiol. 2019;5(2):162–170. doi:10.1016/j.jacep.2018.12.008

26. Boveda S, Higuera L, Longacre C, et al. Two-year outcomes of leadless vs. transvenous single-chamber ventricular pacemaker in high-risk subgroups. Eur Eur Pacing Arrhythm Card Electrophysiol J Work Groups Card Pacing Arrhythm Card Cell Electrophysiol Eur Soc Cardiol. 2023;25(3):1041–1050. doi:10.1093/europace/euad016

27. Saleem-Talib S, Hoevenaars CPR, Molitor N, et al. Leadless pacing: a comprehensive review. Eur Heart J. Published online March 19, 2025:ehaf119. doi:10.1093/eurheartj/ehaf119

28. Kassab K, Patel J, Feseha H, Kaynak E. MICRA AV implantation after transcatheter aortic valve replacement. Cardiovasc Revascularization Med Mol Interv. 2024;63:31–35. doi:10.1016/j.carrev.2024.01.005

29. Shantha G, Brock J, Singleton M, et al. Anatomical location of leadless pacemaker and the risk of pacing-induced cardiomyopathy. J Cardiovasc Electrophysiol. 2023;34(6):1418–1426. doi:10.1111/jce.15925

30. Pipilas D, Frankel DS, Khurshid S. Pacing-induced cardiomyopathy after leadless pacemaker implant: It’s all about location, location, location. J Cardiovasc Electrophysiol. 2023;34(6):1427–1430. doi:10.1111/jce.15944

31. Garweg C, Duchenne J, Vandenberk B, et al. Evolution of ventricular and valve function in patients with right ventricular pacing - A randomized controlled trial comparing leadless and conventional pacing. Pacing Clin Electrophysiol PACE. 2023;46(12):1455–1464. doi:10.1111/pace.14870

32. Haeberlin A, Bartkowiak J, Brugger N, et al. Evolution of tricuspid valve regurgitation after implantation of a leadless pacemaker: A single center experience, systematic review, and meta-analysis. J Cardiovasc Electrophysiol. 2022;33(7):1617–1627. doi:10.1111/jce.15565

33. Andreas M, Burri H, Praz F, et al. Tricuspid valve disease and cardiac implantable electronic devices. Eur Heart J. 2024;45(5):346–365. doi:10.1093/eurheartj/ehad783

34. Sweeney MO, Hellkamp AS, Ellenbogen KA, et al. Adverse effect of ventricular pacing on heart failure and atrial fibrillation among patients with normal baseline QRS duration in a clinical trial of pacemaker therapy for sinus node dysfunction. Circulation. 2003;107(23):2932–2937. doi:10.1161/01.CIR.0000072769.17295.B1

35. Sweeney MO, Prinzen FW. A New Paradigm for Physiologic Ventricular Pacing. JACC. 2006;47(2):282–288. doi:10.1016/j.jacc.2005.09.029

36. Mikhael F. El-Chami, El-Chami MF, Robert C. Kowal, et al. Impact of operator experience and training strategy on procedural outcomes with leadless pacing: Insights from the Micra Transcatheter Pacing Study. Pacing Clin Electrophysiol. 2017;40(7):834–842. doi:10.1111/pace.13094

37. Vora AN, Freeman JV, Enriquez AD. Cutting the Cord: Time for Leadless Pacemakers to Untether Post-TAVR Patients? JACC Cardiovasc Interv. 2024;17(15):1792–1794. doi:10.1016/j.jcin.2024.06.019

38. Mikhael F. El-Chami, El-Chami MF, Matthew Bonner, et al. Leadless pacemakers reduce risk of device-related infection: Review of the potential mechanisms. Heart Rhythm. 2020;17(8):1393–1397. doi:10.1016/j.hrthm.2020.03.019

39. Doshi RN, Ip JE, Defaye P, et al. Dual-chamber leadless pacemaker implant procedural outcomes: Insights from the AVEIR DR i2i study. Heart Rhythm. 2025;0(0). doi:10.1016/j.hrthm.2025.03.1941

40. Reddy VY, Nair DG, Doshi SK, et al. First-in-human study of a leadless pacemaker system for left bundle branch area pacing. Heart Rhythm. 2025;0(0). doi:10.1016/j.hrthm.2025.04.030

